# COVID-19 in patients hospitalized and healthcare workers: what have changed after the first wave in a university hospital

**DOI:** 10.1101/2021.05.11.21257011

**Authors:** Luiz Vinicius Leão Moreira, Gabriela Rodrigues Barbosa, Luciano Kleber de Souza Luna, Alberto Fernando Oliveira Justo, Ana Paula Cunha Chaves, Danielle Dias Conte, Joseane Mayara Almeida Carvalho, Ana Helena Perosa, Klinger Soares Faico Filho, Clarice Neves Camargo, Nancy Bellei

## Abstract

**Objective:** To assess the COVID-19 frequency rates in hospitalized patients (HP) and healthcare workers (HCW), viral load inference, and the impact of vaccination and variants of concern (VOC) during the first pandemic wave.

**Methods:** We evaluated the COVID-19 diagnostics at Hospital São Paulo, Brazil, from March 2020 to April 2021, in 10,202 samples (6,502 HP and 3,700 HCW) tested by RT-qPCR, inferring viral load by cycle threshold (Ct) values, and frequency rates.

**Results:** SARS-CoV-2 was detected in 31.27% of individuals (32.23% HP and 29.80% HCW). The mean age of HP positives was 57.26 ± 18.29 years (median = 59), with a mean Ct value of 25.55 ± 6.07. Neither age nor Ct values in both groups have significantly differed during the first and second waves or even since the predominance of VOC P.1 on March 2021.

**Conclusions:** The COVID-19 epidemic curves of HP and HCW accompanied the variations reported in São Paulo city, as well as the variation of hospitalization and occupancy of ICU beds. The VOC P.1 has no impact on the viral load, since its predominance in March 2021. The vaccination of HCW may have contributed to a decrease in the positivity rates, although more studies will provide a better understanding of the impact of immunization on the COVID-19 pandemic.

## Introduction

Coronavirus disease (COVID-19) has reached over 157 million cases around the world and more than 3.2 million related deaths as of 10 May 2021, according to the World Health Organization (WHO) ^1^. In Brazil, the first COVID-19 case was notified on February 26 in the city of São Paulo. As the world is facing the second wave, Brazil is the third most affected country, with more than 15.1 million confirmed cases and 421 thousand deaths ^1^.

The increase in the number of COVID-19 cases during the second wave is likely to be driven by the emerging new variants of the severe acute respiratory syndrome-related coronavirus 2 (SARS-CoV-2) ^2^. The Variant of Concern (VOC) P.1, first identified in Brazil in January 2021, in Manaus, Northern region of Brazil, has already spread throughout Brazil ^3,4^. Although the lack of evidence between VOC P.1 and disease severity, its infection is being related to an increasing in transmissibility and consequently, a rise in the number of cases ^5^.

The current pandemic constantly threatens the health care system worldwide to collapse ^6^. The handling of a massive number of patients suspected of COVD-19 yielded a great burden for healthcare workers (HCW) and revealed the fragility of the global medical supply chain ^7^.

The COVID-19 epidemic curves provide reliable data to better understand the variation of positivity rates in the groups of HCW and hospitalized patients (HP) and could be used as an indicator of the SARS-CoV-2 infection in the general community, allowing an estimative of the inpatient beds occupied by COVID-19 in public hospitals ^8^. In this sense, we aimed to assess the COVID-19 frequency rates in a university hospital comparing HP and HCW, along with age distribution, indirect viral load inference, the eventual changes during disease course after the first pandemic wave, and the impact of vaccination and emerging VOCs.

## Methods

We assessed the data regarding COVID-19 diagnostics performed at the São Paulo university hospital, Brazil, from March 2020 to April 2021. The study was conducted in compliance with institutional guidelines, approved by the Ethics Committee of São Paulo Federal University (CEP/UNIFESP n. 29407720.4.0000.5505).

A total of 10,202 samples were collected from 6,502 HP and 3,700 HCW including physicians, nurses, resident physicians, logistical support and security staff.

Nasopharyngeal and oropharyngeal swabs were collected in 2.0□mL of sterile lactated Ringer’s solution from all patients admitted at Hospital São Paulo, as well as HCW. The HP presented suggestive Severe Acute Respiratory Syndrome (SARS) symptoms, such as fever, cough, and dyspnea, while HCW presented Influenza-like illness. In some particular cases, other clinical samples such as stool, tracheal aspirate, and other corporal fluids (saliva, urine, serum, and ascitic liquid) were collected to increase diagnosis sensitivity. We also assessed the results of COVID-19 diagnostics required for transferring patients from restricted COVID-19 wards, or intensive care units (ICU), to the non-COVID-19 ones, such as private, dialysis, or chemotherapy rooms.

For RNA preparation, the samples were homogenized and purified using the Quick-RNA Virus kit (Zymo Research, USA) according to the manufacturer’s instructions. Molecular detection was performed by reverse transcription, real-time polymerase chain reaction (RT-qPCR) assay, using the GeneFinder COVID-19 Plus RealAmp Kit (OSANG Healthcare, Korea), which targets the E (envelope), N (nucleocapsid), RdRp (RNA-dependent RNA Polymerase) genes of SARS-CoV-2, and human ribonuclease P (RNase P) as an internal control. A positive result was considered with a Ct (cycle threshold) value ≤ 40, for at least two SARS-CoV-2 genes (RdRp, E, and N), following the manufacture’s results interpretation.

We analyzed the RT-qPCR Ct values as a semi-quantitative parameter, in order to infer viral load, meaning that the lower the value, the higher is the viral load. The lowest Ct values of individuals were computed from July 2020, when we were certified by the Laboratory Platform for Diagnosis of Coronavirus in Sao Paulo, coordinated by Butantan Institute, to April 2021.

### Statistical Analysis

Frequency rates were analyzed using the chi-squared test. Age and Ct values were compared by the Mann-Whitney test or ANOVA with Tukey’s multiple comparisons test. Statistical differences were considered for *p* < 0.05. The values were expressed as mean ± standard deviation. All analyses were performed using the software GraphPad Prism v.6.01.

## Results

Clinical samples were almost comprised of nasopharyngeal and oropharyngeal swabs (95.75%), and also tracheal aspirate (2.05%), stool (1.67%), and other fluids (0.53%). We excluded the entry of more than two samples from the same individual per month to avoid bias on the frequency rates. Hence, 8,644 were included in the analysis (5,241 samples from HP, and 3,403 from HCW).

From March 2020 to April 2021, we performed a mean of 617.43 ± 187.05 COVID-19 diagnostics per month (374.36 ± 93.73 for HP, and 243.07 ± 128.44 for HCW). The overall SARS-CoV-2 frequency rate was 31.27% (2703/8644). In HP and HCW, the frequency rates were 32.23% (1689/5241), and 29.80% (1014/3403), respectively, which have significantly differed (*p* = 0.0173). The frequency rates of HP and HCW by month per year, and their statistical differences, are shown in Table 1.

**Table 1:** Frequency rates of SARS-CoV-2 in hospitalized patients and healthcare workers by year/month, from March 2020 to April 2021.

| Year/Month | HP |  | HCW |  | *P-value* |
| --- | --- | --- | --- | --- | --- |
|  | samples | positives<br>n (%) | samples | positives<br>n (%) |  |
| 2020 |  |  |  |  |  |
| March | 209 | 54 (25.84) | 119 | 39 (32.77) | 0.1802 |
| April | 335 | 173 (51.64) | 467 | 173 (37.04) | < 0.0001* |
| May | 486 | 237 (48.77) | 354 | 115 (32.49) | < 0.0001* |
| June | 469 | 205 (43.71) | 515 | 264 (51.26) | 0.0178* |
| July | 443 | 155 (34.99) | 307 | 102 (33.22) | 0.6167 |
| August | 346 | 95 (27.46) | 174 | 35 (20.11) | 0.0681 |
| September | 290 | 57 (19.66) | 173 | 30 (17.34) | 0.5375 |
| October | 309 | 34 (11.00) | 122 | 17 (13.93) | 0.3970 |
| November | 334 | 60 (17.96) | 255 | 51 (20.00) | 0.5313 |
| December | 303 | 91 (30.03) | 212 | 36 (16.98) | 0.0007* |
| 2021 |  |  |  |  |  |
| January | 367 | 146 (39.78) | 255 | 62 (24.31) | < 0.0001* |
| February | 356 | 91 (25.56) | 105 | 24 (22.86) | 0.5735 |
| March | 567 | 195 (34.39) | 217 | 48 (22.12) | 0.0009* |
| April | 427 | 96 (22.48) | 128 | 18 (14.06) | 0.0386* |
| *Total* | *5,241* | *1,689 (32.23)* | *3,403* | *1,014 (29.80)* | *0.0173\** |

HP, hospitalized patients. HCW, healthcare workers.
\*Significant at $p < 0.05$

Higher frequency rates of positive samples in HP were detected in adults aging 50-59 years (42.11%), and elderly over 60 years old (43.03%), with a significant difference from those of HCW (30.64%, *p* < 0.0001; and 30.51%, *p* = 0.0076, respectively) for the same age groups. On the other hand, in HCW, the frequency rates have not differed statistically within age groups.

The epidemic curves of COVID-19 observed at Hospital Sao Paulo for HP and HCW were comparable most of the time during the course of disease over time (Figure 1), although the SARS-CoV-2 frequency rates in HP were significantly higher than HCW in April, May, and December 2020, and January, March and April 2021, and the opposite in June 2020 (Table 1).

**Figure 1:**
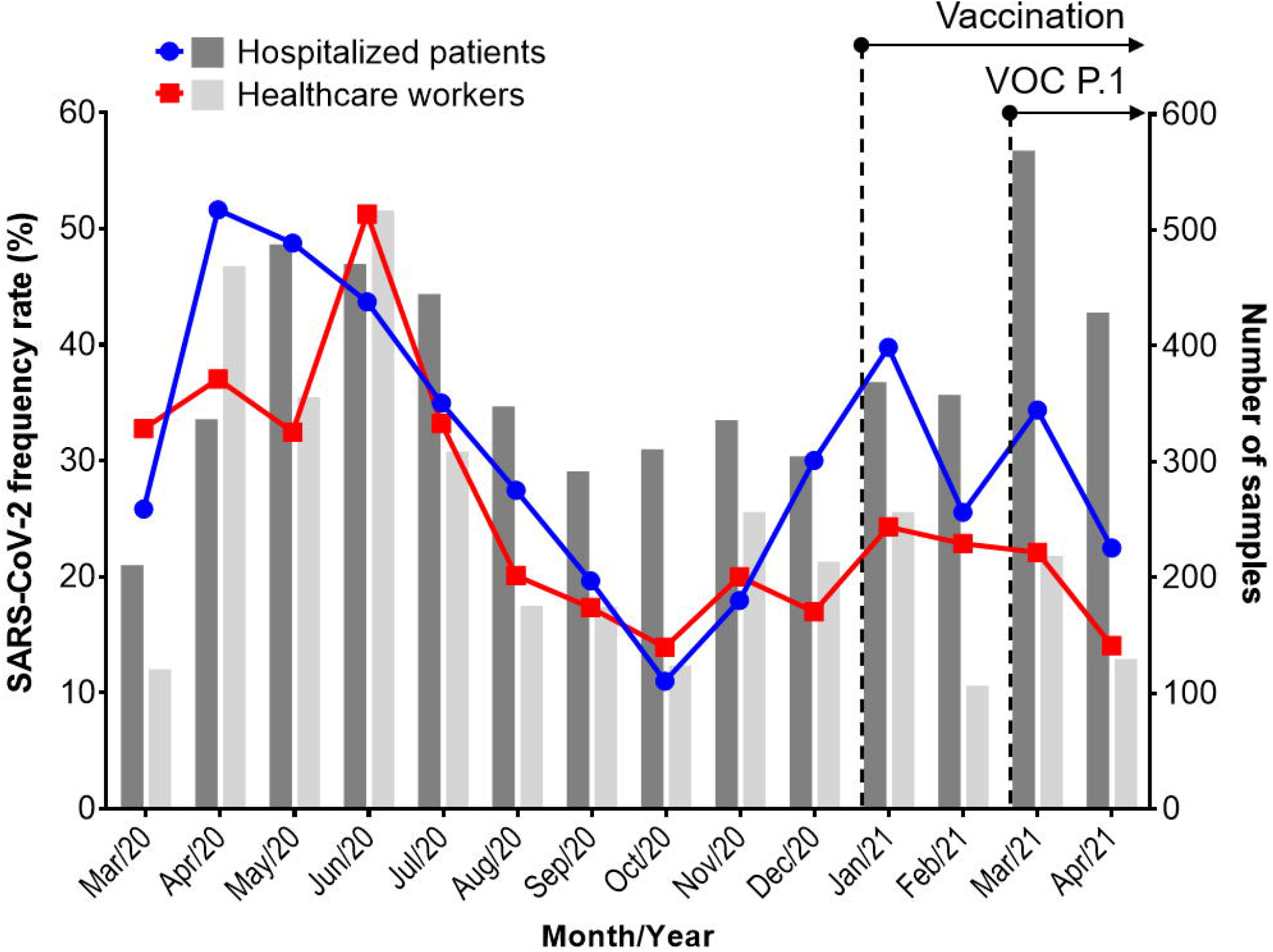
SARS-CoV-2 frequency rates (left y-axis, blue and red lines) and the number of samples analyzed (right y-axis, gray bars). Dotted lines and arrows indicate the beginning of the SARS-CoV-2 vaccination campaign and detection of VOC P.1 at Hospital São Paulo.

During the first SARS-CoV-2 wave, from March to October 2020, the positivity in HP and HCW reached their peaks in April (51.6%) and June (51.2%), respectively, whereas the lowest frequency rates were observed in October for both groups, with 13.9% for HP and 11.00% for HCW (Table1, Figure 1). The frequency rates started to increase again in November 2020, reaching a new peak in January 2021, when HCW started to be immunized in Brazil.

In December 2020, January and April 2021, frequency rates of SARS-CoV-2 were significantly higher in HP in relation to HCW (*p* = 0.0007, *p* < 0.0001, *p* = 0.0176, respectively). Interestingly, frequency rates in HCW were almost the same during the first three months of 2021 (24.31%, 22.86%, and 22.12%, respectively) and dropped to 14.06% in April. In 2021, for HP, the frequency rate oscillated in February (25.56%), and March (34.39%), and dropped in April (22.48%).

In March 2021 we tested the highest number of HP samples since the beginning of the COVID-19 pandemic when VOC P.1 was first confirmed at Hospital São Paulo in more than 84% of all sequenced samples ^9^.

The age of HP ranged from 0 to 101 years, with a mean of 47.28 ± 24.56 years, and a median of 52 years. On the HP positive for SARS-CoV-2, the age range was the same, with a mean of 57.26 ± 18.29 years, and a median of 59 years (range 55-66) over the analyzed period. There was a significant difference (*p* = 0.0023) between the months with the lowest (52.75 ± 20.18) and highest (62.99 ± 18.62) mean ages, which corresponded to November and December 2020, respectively (Figure 2). There was no significant difference in the mean ages since the predominance of VOC P.1 on March 2021.

**Figure 2:**
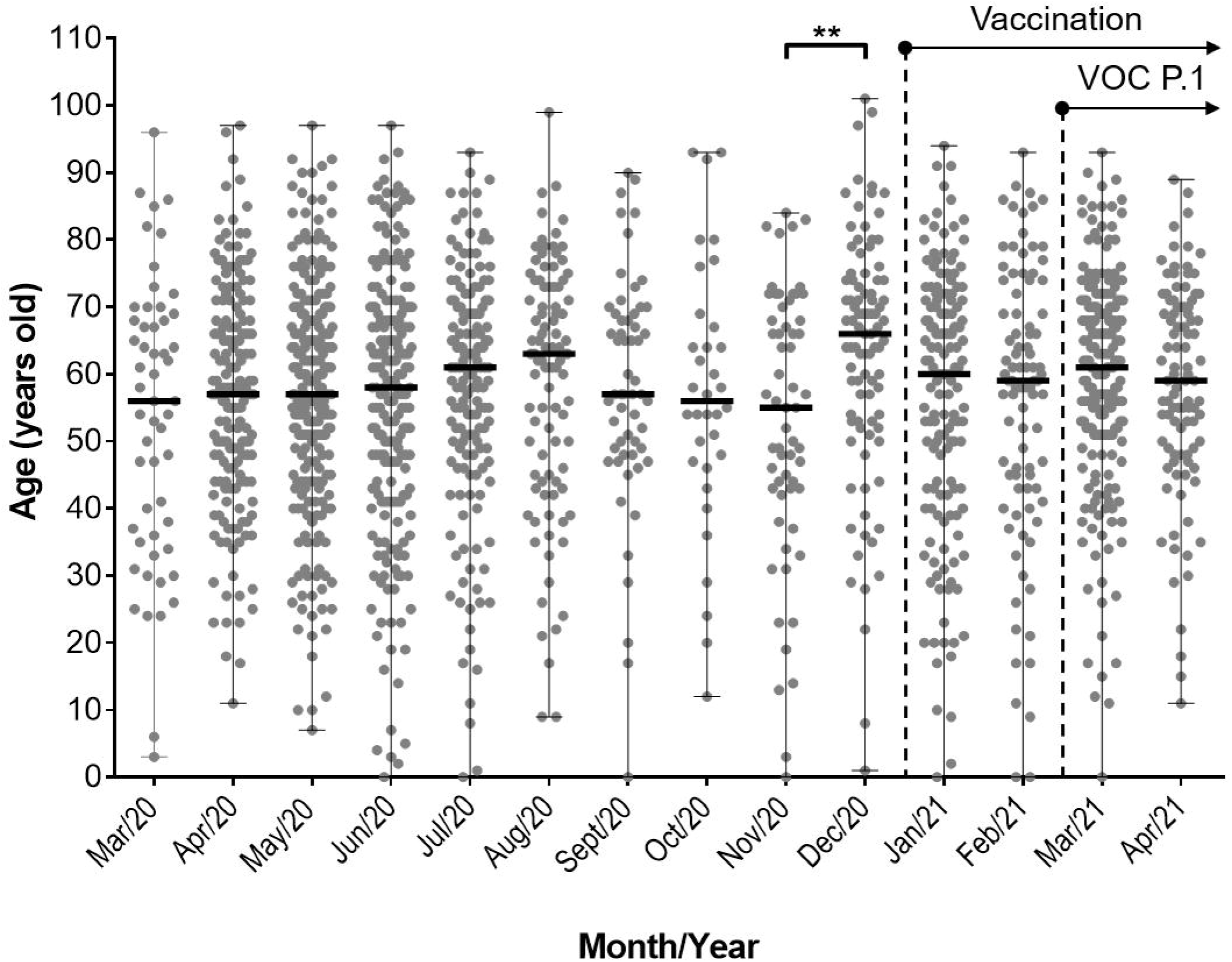
Monthly age distribution of hospitalized patients positive for SARS-CoV-2. Upper and lower bars indicate maximum and minimum values, respectively. Horizontal bars indicate median values. Dotted lines and arrows indicate the beginning of the SARS-CoV-2 vaccination campaign and detection of VOC P.1 at Hospital São Paulo. *significant at *p* < 0.05.

For HCW, the age varied from 14 to 92 years (mean = 37.24 ± 11.4; median = 36), and in the SARS-CoV-2 positives, the age varied from 19 to 78 years (mean = 37.58 ± 11.49; median = 36), with no statistical differences within the mean age over the time. The RT-qPCR Ct values, computed from July 2020 to April 2021, ranged from 9 to 40 (mean = 25.55 ± 6.07; median = 25) for HP, and 10 to 40 (mean = 23.68 ± 5.82; median = 23) for HCW, with a significant difference between the groups (*p* = 0.0023). The mean Ct values have not statistically differed within HP and HCW groups over time, even since the beginning of HCW vaccination or VOC P.1 predominance (Figure 3).

**Figure 3:**
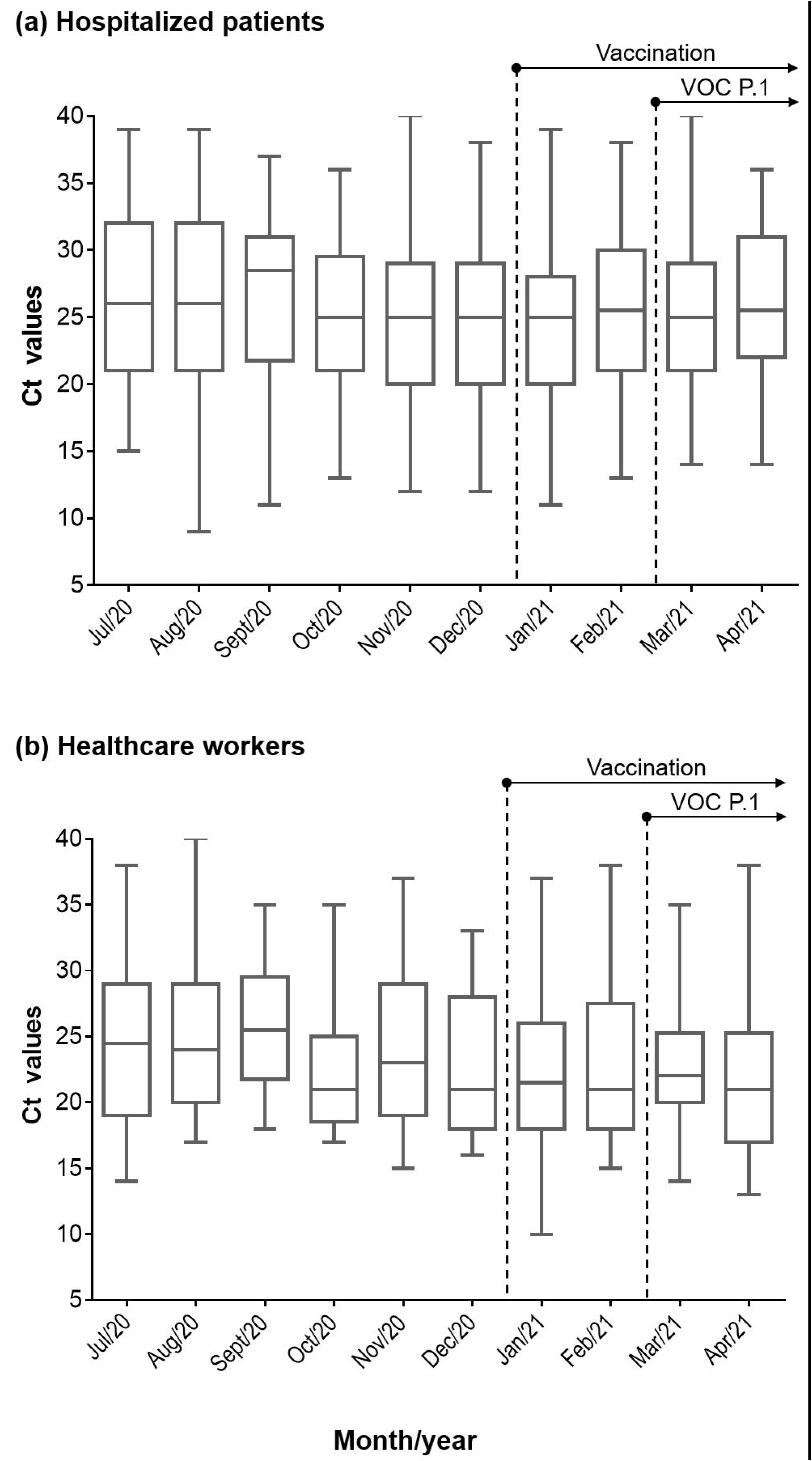
Distribution of RT-qPCR Ct values from May 2020 to March 2021. (a) Hospitalized patients. (b) Healthcare workers. Upper and lower bars indicate maximum and minimum values. Horizontal bars within boxes indicate median values. Boxes extend from the 25^th^ to 75^th^ percentiles of Ct values. Dotted lines and arrows indicate the beginning of the SARS-CoV-2 vaccination campaign and detection of VOC P.1 at Hospital São Paulo.

## Discussion

In this study, we could assess the COVID-19 infection in HP and HCW over the first wave, and the beginning of the second, at a university hospital. The variation observed of COVID-19 cases in the epidemic curves of HP and HCW accompanied the variations reported by the municipal health department of São Paulo city ^10^. The official database of the city reported a peak in the number of confirmed COVID-19 cases for HP in May 2020, as well as an increased occupancy of ICU beds in public and private services. After this critical period, there was a decrease in confirmed cases. However, in January 2021, the number of cases started to rise again, reaching virtually the total occupancy of ICU beds all over the city.

As the world faces the second wave of SARS-CoV-2, the emerging new variants are often being associated with increased transmissibility. As we have shown, the positivity rates started to rise again in November 2020. Notably, the VOC P.1 was first detected in January 2021 and was responsible for the second wave in Manaus (Northern region of Brazil) since late 2020. Recently, a SARS-CoV-2 whole-genome survey at Hospital São Paulo confirmed that 84.4% of sequenced samples from the first two weeks of March 2021 were VOC P.1, demonstrating that it has already spread throughout the country ^9^. The higher frequency rates were observed in HP, especially in patients over 50 years of age, who were responsible for more than 70% of confirmed COVID-19 cases. These results should be explained by the fact that comorbidities tend to increase among individuals above this age, as well as declining immunity ^11^. However, the mean age of HP has not significantly differed since March 2021, when we observed the predominance of VOC P.1, in relation to the first wave.

Virtually, all molecular detection of SASRS-CoV-2 was made from naso- or oropharyngeal swabs. In patients with negative results but with clinical presentation suggestive of COVID-19, new swabs were collected together with other clinical samples, according to availability, such as tracheal aspirates, and stool, which helped to increase diagnostic sensitivity ^12^.

The inferred mean viral load (Ct values) has not shown a significant difference within HP and HCW over time, although the viral load in HCW was significantly higher than that observed in HP. This can be explained by the fact that the detection of SARS-CoV-2 in HCW was performed during the first days of symptoms, usually up to the third day. As higher viral loads have been associated with more COVID-19 severe cases and death in São Paulo ^13^, an increase in viral loads associated with VOC P.1 infection would be expected, regarding greater pathogenicity, which was not observed since at least its predominance in March 2021 ^9^.

The emerging SARS-CoV-2 variants and the immunization of the population are major events through the second wave of COVID-19. Although the rapid spread of VOC P.1 is not strictly related to an increase in severity or death, it is likely to be responsible for an increase of transmissibility which resulted in a rise in hospitalization and ICU occupancy.

The vaccination of HCW may have contributed to a decrease in the positivity rates. However, vaccine effectiveness against SARS-CoV-2, especially on VOCs such as P.1, can only be evaluated when the majority of the population will already have been immunized. Nonetheless, a preliminary study has shown that VOC P.1 may escape neutralizing antibodies from plasma of convalescent COVID-19 patients and sera from individuals vaccinated with Pfizer/BioNTech vaccine ^14^. It is noteworthy that the vaccination of HCW in São Paulo started on 19 January 2021 regardless of the age group. In the general community, the vaccination campaign was initiated on 18 February 2021, by age group, starting with individuals aging > 90 years of age, and until 05 April attended the public over 60 years old ^15^. At this pace, individuals over 50 years old, responsible for more than 70% of confirmed COVID-19 cases, should be vaccinated until June 2021. In this sense, more epidemiologic surveillance studies with the vaccinated population will provide a better understanding of the impact of immunization on the COVID-19 pandemic and the circulation of novel variants.

## Data Availability

Not Avaliable

## Acknowledgements

L.V.L.M, L.K.S.L, A.P.C.C., D.D.C., and J.M.A.C. are fellows of the Coordenação de Aperfeiçoamento de Pessoal de Nível Superior (CAPES). G.R.B. is supported by grant 2020/11719-0 São Paulo Research Foundation (FAPESP). A.F.O.J. is supported by grant 27968 Fundação de Desenvolvimento da Pesquisa (FUNDEP; FINEP, REDE COVID).

## References

1. WHO. WHO Coronavirus (COVID-19) Dashboard. 2021. https://covid19.who.int/, 2021.

2. Esper FP, Cheng YW, Adhikari TM, et al. Genomic Epidemiology of SARS-CoV-2 Infection During the Initial Pandemic Wave and Association With Disease Severity. JAMA Netw Open 2021;4:e217746.

3. Faria NR, Claro IM, Candido D, et al. Genomic characterisation of an emergent SARS-CoV-2 lineage in Manaus: preliminary findings. 2021. https://virological.org/t/genomic-characterisation-of-an-emergent-sars-cov-2-lineage-in-manaus-preliminary-findings/586.

4. Resende PC, Graf T, Paixao ACD, et al. A Potential SARS-CoV-2 Variant of Interest (VOI) Harboring Mutation E484K in the Spike Protein Was Identified within Lineage B.1.1.33 Circulating in Brazil. Viruses 2021;13.

5. Guo S, Liu K, Zheng J. The Genetic Variant of SARS-CoV-2: would It Matter for Controlling the Devastating Pandemic? Int J Biol Sci 2021;17:1476–1485.

6. Adams JG, Walls RM. Supporting the Health Care Workforce During the COVID-19 Global Epidemic. JAMA 2020;323:1439–1440.

7. Ranney ML, Griffeth V, Jha AK. Critical Supply Shortages - The Need for Ventilators and Personal Protective Equipment during the Covid-19 Pandemic. N Engl J Med 2020;382:e41.

8. Faico-Filho KS, Carvalho JMA, Conte DD, de Souza Luna LK, Bellei N. COVID-19 in health care workers in a university hospital during the quarantine in Sao Paulo city. Braz J Infect Dis 2020;24:462–465.

9. Barbosa GR, Moreira LVL, Justo AFO, et al. Rapid spread and high impact of the Variant of Concern P.1 in the largest city of Brazil. J Infect 2021.

10. SMS. Inquérito Sorológico Adultos (> 18) para SARS-CoV-2 2021: Evolução da prevalência da infecção no MSP. 2021. https://www.prefeitura.sp.gov.br/cidade/secretarias/upload/saude/2021_03_12_inq_soro_2021_fase_4_pdf.pdf.

11. Bhatla A, Mayer MM, Adusumalli S, et al. COVID-19 and cardiac arrhythmias. Heart Rhythm 2020;17:1439–1444.

12. Moreira LVL, de Souza Luna LK, Barbosa GR, et al. Test on stool samples improves the diagnosis of hospitalized patients: Detection of SARS-CoV-2 genomic and subgenomic RNA. J Infect 2021;82:186–230.

13. Faico-Filho KS, Passarelli VC, Bellei N. Is Higher Viral Load in SARS-CoV-2 Associated with Death? Am J Trop Med Hyg 2020;103:2019–2021.

14. Hoffmann M, Arora P, Gross R, et al. SARS-CoV-2 variants B.1.351 and P.1 escape from neutralizing antibodies. Cell 2021;184:2384–2393 e2312.

15. SMS. Vacinação contra COVID-19 no município de São Paulo. 2021. https://www.prefeitura.sp.gov.br/cidade/secretarias/saude/vigilancia_em_saude/doencas_e_agravos/coronavirus/index.php?p=307599.

